# Rapid initiation of nasal saline irrigation to reduce severity in high-risk COVID+ outpatients: a randomized clinical trial compared to a national dataset observational arm

**DOI:** 10.1101/2021.08.16.21262044

**Authors:** Amy L. Baxter, Kyle R. Schwartz, Ryan W. Johnson, Ann-Marie Kuchinski, Kevin M. Swartout, Arni S. R. Srinivasa Rao, Robert W. Gibson, Erica Cherian, Taylor Giller, Houlton Boomer, Matthew Lyon, Richard Schwartz

## Abstract

**Importance:** SARS-CoV-2 enters the nasopharynx to replicate; nasal irrigation soon after diagnosis could reduce viral load and inhibit furin cleavage necessary for cell entry, thereby reducing morbidity and mortality.

**Objective:** To determine whether initiating nasal irrigation after COVID-19 diagnosis reduces hospitalizations and death in high-risk outpatients, and whether irrigant composition impacts severity.

**Design:** Unblinded randomized clinical trial of two nasal irrigation protocols in older outpatients PCR positive for SARS-CoV-2, with an observational arm using laboratory-confirmed cases in the CDC COVID-19 Case Surveillance dataset.

**Setting:** Single-lab community testing facility associated with the emergency department (ED) in Augusta, GA.

**Participants:** A consecutive sample of high-risk adults were enrolled within 24 hours of a positive COVID-19 test between September 24 and December 21 of 2020. Patients aged 55 and older were remotely consented. Among 826 screened, 321 of 694 eligible patients were unable to be reached, 294 refused participation, and 79 participants were enrolled.

**Interventions:** Participants were randomly assigned adding 2.5 mL povidone-iodine 10% or 2.5 mL sodium bicarbonate to 240 mL of isotonic nasal irrigation twice daily for 14 days.

**Main Outcomes and Measures:** The primary outcome was hospitalization or death from COVID-19 within 28 days of enrollment by daily self-report confirmed with phone calls and hospital records, compared to the CDC Surveillance Dataset covering the same time. Secondary outcomes compared symptom resolution by irrigant additive.

**Results:** Seventy-nine high-risk participants were enrolled (mean [SD] age, 64 [8] years; 36 [46%] women; 71% Non-Hispanic White), with mean BMI 30.3. Analyzed by intention-to-treat, by day 28, COVID-19 symptoms resulted in one ED visit and no hospitalizations in 42 irrigating with alkalinization, one hospitalization of 37 in the povidone-iodine group, (1.27%) and no deaths. Of nearly three million CDC cases, 9.47% were known to be hospitalized, with an additional 1.5% mortality in those without hospitalization data. The total risk of hospitalization or death (11%) was 8.57 times that of enrolled patients (SE=2.74; P=.006). 62 completed daily surveys (78%), averaging 1.8 irrigations/day. Eleven had irrigation complaints, and four discontinued. There were no significant differences by additive.

**Conclusion:** SARS-CoV-2+ participants initiating nasal irrigation were over 8 times less likely to be hospitalized than the national rate.

**Trial Registration:** ClinicalTrial.gov Identifier: NCT04559035

**Author Approval:** All authors have filled out ICMJE and approved submission.

**Conflict of Interest Statement:** Materials were provided by Neilmed Inc. and Rhinosystems Inc. The study was supported by funding from the Bernard and Anne Gray Donor Advised Fund Community Foundation for Greater Atlanta, Neilmed Inc., and Rhinosystems. No authors have conflict of interest.

**Key Points:** *Question:* After testing positive for COVID-19, will rapidly initiating nasal irrigation reduce the risk of morbidity and mortality compared to a national dataset?

*Findings:* A consecutive sample of 79 high-risk adults (mean age 64, BMI 30.3) were randomized to initiate one of two nasal irrigation protocols within 24 hours of a positive COVID-19 test. Compared to a CDC COVID-19 National Dataset observational arm, 1.27% of participants initiating twice daily nasal irrigation were hospitalized or died, compared to 11%, a significant difference.

*Meaning:* In high-risk outpatients testing positive for SARS-CoV-2 who initiated nasal irrigation rapidly after diagnosis, risk of hospitalization or death was eight times lower than national rates reported by the CDC.

## Background and Objectives

Pharmacologic and immunologic COVID-19 therapeutics seek to inhibit mechanical binding of the SARS-CoV-2 spike protein-receptor to the ACE2 receptor, and spike segment furin cleavage necessary for cell entry. Sungnak et al. localized the necessary co-expression of ACE2 and protease TMPRSS2 primarily in the ciliated nasal epithelia,^1^ supporting the clinical correlation of nasal viral load and severity and suggesting a location for early intervention. The increased infectiousness resulting from physical changes with viral spike protein mutations support a mechanical opportunity to interrupt viral particle receptor binding and entry.^2^ The observation that saline can inhibit furin cleavage^3^ suggests a mechanical therapeutic option – nasal irrigation – may be particularly effective against this pathogen.

Nasal irrigation under pressure, or “nasal lavage”, has been demonstrated to safely reduce the duration and severity of both *Coronaviridae* and illnesses like flu with shorter incubation periods.^4-7^ Repeated irrigation should be most effective for pathogens with prolonged incubation, local non-hematogenous spread, and variolation where viral load impacts severity.

Given research supporting the virucidal activity of povidone-iodine against MERS and SARS-CoV-2^8-10^ and the possible impact of alkalinization to reduce SARS-CoV-1 viral cell fusion and entry,^11^ patients were randomized to add alkalinization or povidone-iodine to pressurized nasal lavage. We hypothesized rapid initiation of nasal irrigation after testing positive would reduce the severity of COVID-19. Our primary outcome was COVID-19 hospitalization or death, with secondary outcomes of symptom duration, severity, and household spread.^12,13^ If clinically effective, irrigation could be an inexpensive option rapidly available worldwide.

## Methods

### Trial Design

The Rapid Initiation of Nasal Saline Irrigation (RINSI) study was an unblinded randomized clinical trial of 240 ml saline nasal irrigation supplemented with 2.5ml of either sodium bicarbonate or 10% povidone iodine in high-risk outpatients aged 55 years and older recently PCR positive for SARS-CoV-2. The comparative observational arm comprised laboratory-confirmed cases in the CDC COVID-19 Case Surveillance Dataset of patients 50 and older during the same time interval.^14^ Primary outcomes were hospitalization or death within 28 days. The trial statistical analysis plan and recruitment protocol appear in Supplement 1. The study was approved by the institutional review board at Augusta University in Augusta, Georgia and was registered at ClinicalTrials.gov NCT04559035. All participants provided remote informed consent in English.

### Study Setting and Recruitment

This trial was conducted in Augusta, Georgia. Patients testing positive for COVID-19 by nasal swab or saliva PCR processed at a single lab at the Augusta University were recruited from September 24, 2020 to December 21, 2020. The 28-day follow-up was completed January 18, 2021. (See Appendix 1). The daily laboratory-generated list of COVID-19 tests was screened for patient age, first positive test in the system, and location within 25 miles of Augusta University. Prospective participants were called consecutively between the hours of 9:00am through the early afternoon five to six days a week. When test results exceeded staffing, the list was randomized for calling order. Participants interested in participation were assessed over the phone for inclusion criteria, and remote informed consent was completed per IRB policy.

Using COVID-19 precautions (masks, maintaining 6-foot physical distance, door drop off), same-day home delivery of materials included a nasal irrigation device with 28+ day supplies, two gallon jugs of distilled water, the consent form, instructions, and the study additive (baking soda or povidone-iodine) with a 2.5 mL scoop. One of two high pressure^15^ irrigation devices (NAVAGE [Rhinosystems Inc.] or Neilmed Sinus Rinse [Neilmed Inc.] was provided, alternating days for each brand.

#### Eligibility Criteria

Participants had to be able to read the informed consent in English, agree to nasal lavage for 14 days with a 14-day follow-up, provide a back-up contact, and receive materials and initiate irrigation that day. Exclusion criteria included current supplemental oxygen therapy, unwillingness to try or current use of nasal irrigation, nasal surgery within the past year or chronic sinusitis, prior COVID-19 infection or positive test, symptoms longer than 7 days, inability to complete surveys by computer or smartphone, and allergies to iodine or shellfish.

#### Randomization

Patients were randomized to rinse with 240cc saline including 0.5 mL 10% povidone-iodine (0.1% final concentration) or 2.5 mL sodium bicarbonate twice daily for 14 days. Randomization was stratified by sex in 10 blocks of 10 random numbers using Random.org. With odd numbers signifying alkaline and even povidone-iodine, numbered opaque envelopes were prepared in separate sequences for male or female participants to be opened after consent.

### Interventions and Masking

Masking of study treatments (alkalinization or 0.1% povidone-iodine) was not undertaken. Study personnel reviewing hospital records to verify admission or death were masked to intervention.

#### Main Outcomes and Measures

The primary outcome was hospitalization or death from COVID-19 within 28 days of enrollment, by self-report, phone calls, and the testing site hospital’s electronic medical record. Secondary outcomes in enrolled patients compared symptom resolution, severity, household spread, adherence to nasal irrigation, and any impact of irrigant additive. Symptoms tracked included loss of smell or taste, fatigue, fever >100.4°F, chills, muscle aches, runny nose, cough (new onset or worsening of chronic cough), shortness of breath, nausea or vomiting, headache, abdominal pain, and diarrhea.

In addition to demographic data, after enrollment patients were asked preexisting medical history as found on the CDC person of interest form, including Obesity, Chronic Lung disease (Emphysema, COPD), Asthma, Type 1 or 2 Diabetes, Cardiovascular Disease, Hypertension, Chronic Renal Disease, Immunocompromised, and weight and height to calculate obesity defined as BMI>30. Prompts were sent to participants via email from Qualtrics twice daily. To verify irrigation, patients uploaded pictures of used irrigation materials. An investigator called the patient or their designated contact at day 2, 7, 14, and 28 to verify irrigation, hospitalization, or answer any questions.

Hospitalization and mortality data were compared to the National CDC Case Surveillance Public Use Dataset.^14^ Twelve elements are shared with the CC for all COVID-19 cases, including date of first positive specimen, report to CDC, illness, and summary “case earliest date”, laboratory-confirmed or suspected, symptom onset, and demographic data. Hospitalization, pre-existing conditions, and mortality data have four options: yes, no, unknown (marked on form), or missing (nothing recorded). Following CDC research recommendations, we matched laboratory confirmed cases by “case earliest date” with our testing dates.

### Statistical Analysis

Chi-square was used to evaluate differences in demographic proportions of sex, race, and age by 10-year tranche. We used an exact binomial test with Clopper-Pearson confidence intervals to compare observed hospital admission rates among participants compared with national rates of severe disease (admission or death) published by CDC. The exact binomial test is well-suited to assess the probability of observing the proportion of patients in this study. (`IBM SPSS Statistics for Windows, Version 27.0. Armonk, NY: IBM Corp`.)

To avoid overestimating hospitalization rates by reporting bias, the denominator included all laboratory-confirmed cases, including when hospitalization status was missing or unreported. In addition to reported hospitalizations, we included deaths in the numerator only for confirmed cases where hospitalization was unknown or missing; did not include deaths in cases where hospitalization status was known to avoid counting outcomes with increased severity twice. As an indicator of the impact of unreported hospitalizations, we report relative risk using both this most conservative denominator (underestimating hospitalizations), and for hospitalizations using only known yes or no responses in the denominator (potential for overestimating due to reporting bias). (MedCalc Software Ltd. https://www.medcalc.org/calc/relative_risk.php (Version 20.009)).

## Results

During the study period, 826 unique patients aged 55 and older who tested positive for COIVD-19 were screened for study eligibility. Of the 694 eligible, 321 were unable to be reached, 294 refused participation, and 79 participants were able to be enrolled and receive irrigation materials on the day of contact (Figure 1).

**Figure 1:**
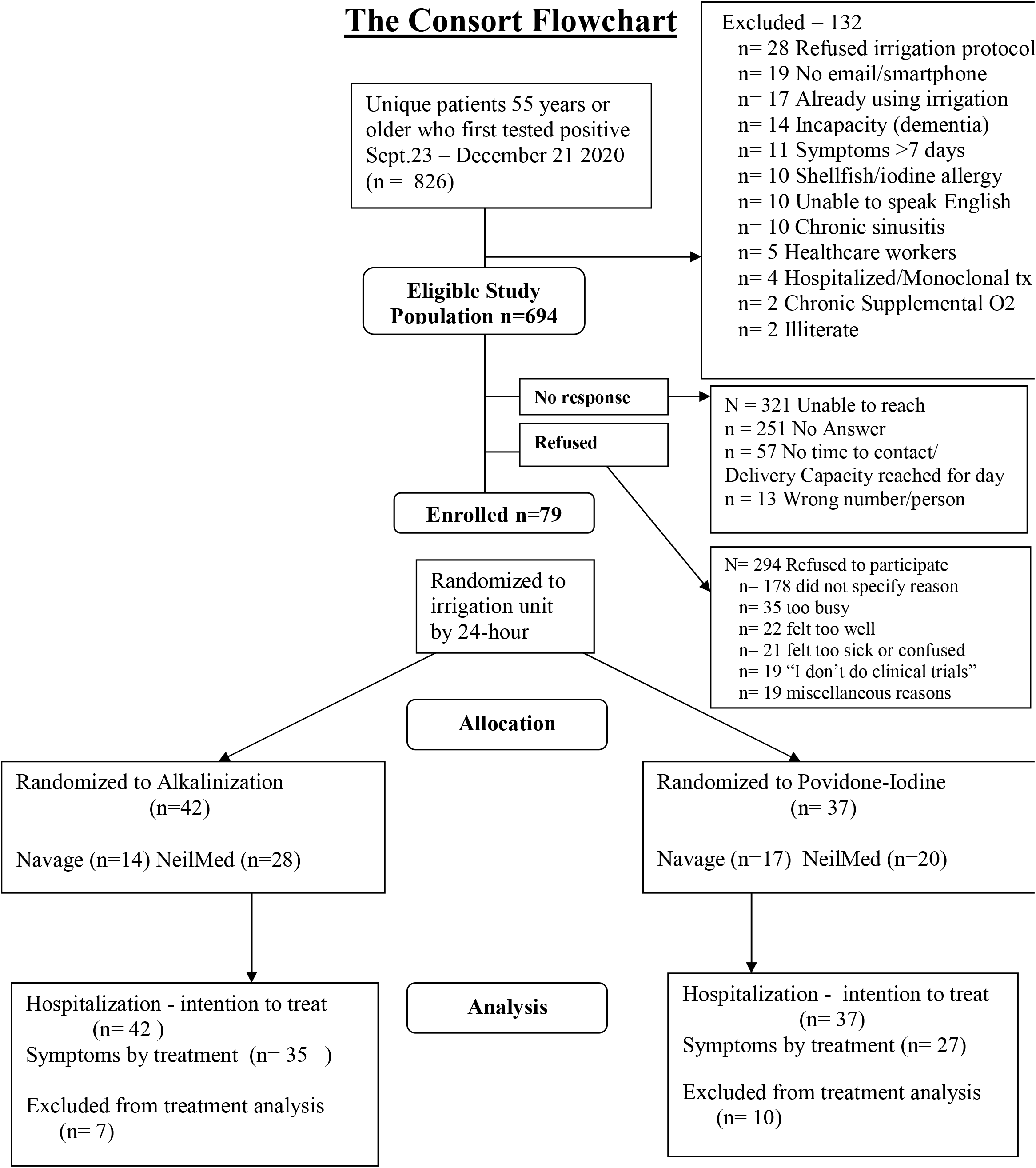
CONSORT diagram

Admissions occurred for 1/37 assigned to povidone-iodine and 0/42 patients in the alkalinization group (1.27%). One patient in the alkalinization group reported a COVID-19 related ED visit without admission (pre-monoclonal antibody availability), one patient reported an ED visit for a minor trauma, and one patient was admitted for a syncopal episode after resolution of COVID symptoms. These events were verified in the EHR database, confirmed with the day 28 phone call, and there were no additional ED visits or hospitalizations found in consented patients.

Between September 23, 2020 and December 21, 2020, for patients 50 years and older, of 2,962,541 laboratory-confirmed cases, 280533 (9.47%) were reported hospitalized. Complete hospitalization information was available for 45%. Where hospitalization status was unknown/missing, 44,773 deaths were reported, or 1.5%. Thus, hospitalization (or death when admission status was unknown/missing) occurred in 11%, 8.57 times the hospital admission rate of nasal irrigation participants (SE=2.74; P=.006), (Figure 2). The relative risk for nasal irrigation subjects was .119 (95%CI .0169 to .833, P=.032). Using only the 1,328,778 cases where hospitalization status was reported, the relative risk of hospitalization was .0594 (95%CI .0085 to 0.416, P=.0045) when using nasal irrigation, with a number needed to treat of 5. For the 1,002,050 CDC cases for whom both hospitalization and death were reported, 9.18% of patients expired.

**Figure 2:**
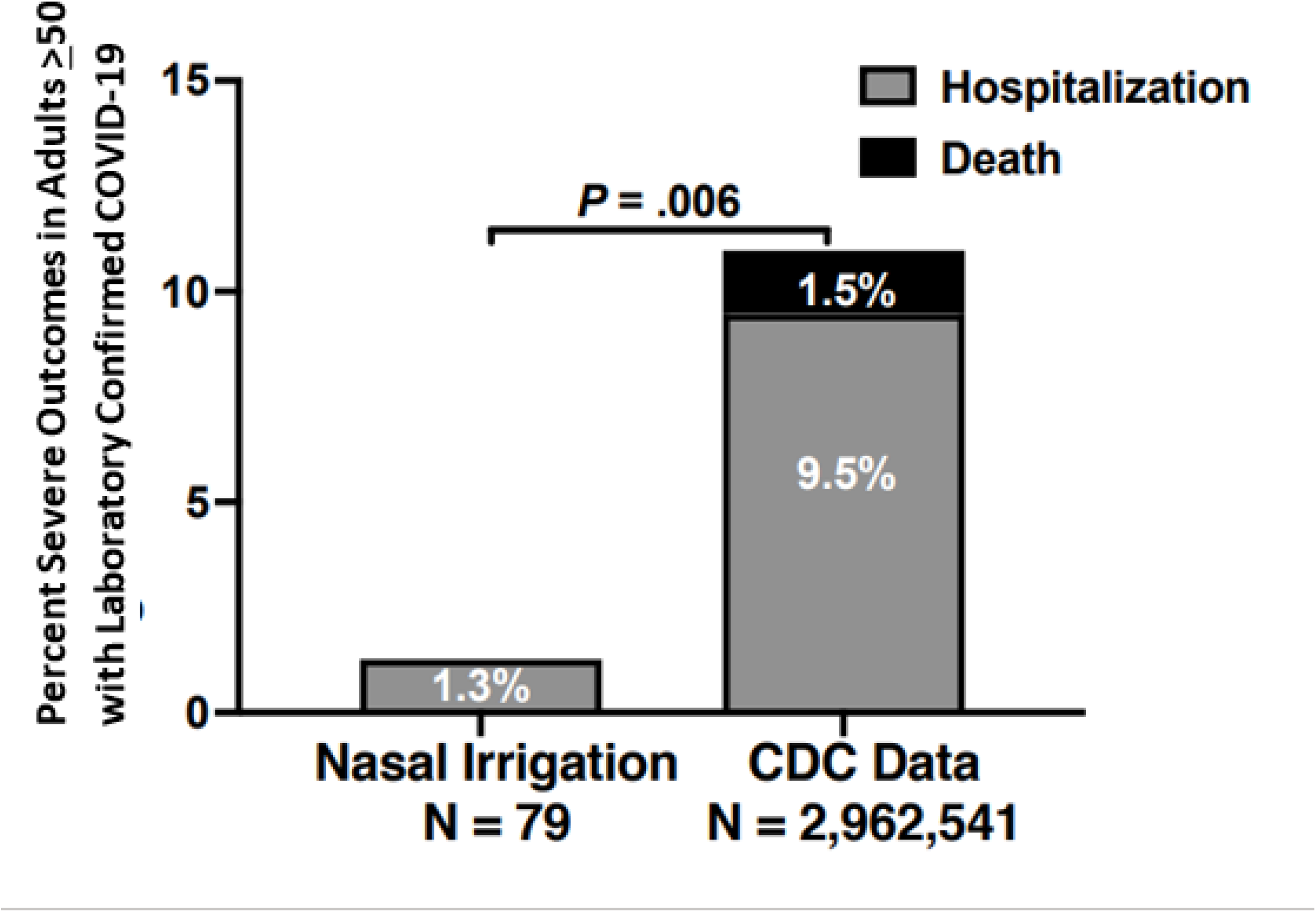
Percent Severe Outcomes in Nasal Irrigation Group Compared to CDC Dataset Percent of patients ≥55 in the prospective nasal irrigation group who were hospitalized compared to the number of patients age ≥50 in the CDC National Dataset reported hospitalized, or with death reported if hospitalization information was not reported or missing.

The CDC dataset only reported race for 65% of confirmed cases; those reported had a lower proportion of minority patients than enrolled irrigation patients (Table 1). There was no difference in age or sex from our population. Of the 79 enrolled, 53 patients completed the initial symptom and history questionnaire. (Table 2) An online daily symptom and irrigation data collection survey was completed by 62 participants (median 12 of 14 days [IQR 5,13.75]). Of enrolled patients, 68% had a pre-existing condition, 45% had multiple conditions, and the average BMI was 30.27. Patients reported a median of 3.3 days [IQR 2,5] of symptoms prior to enrollment.

**Table 1:**
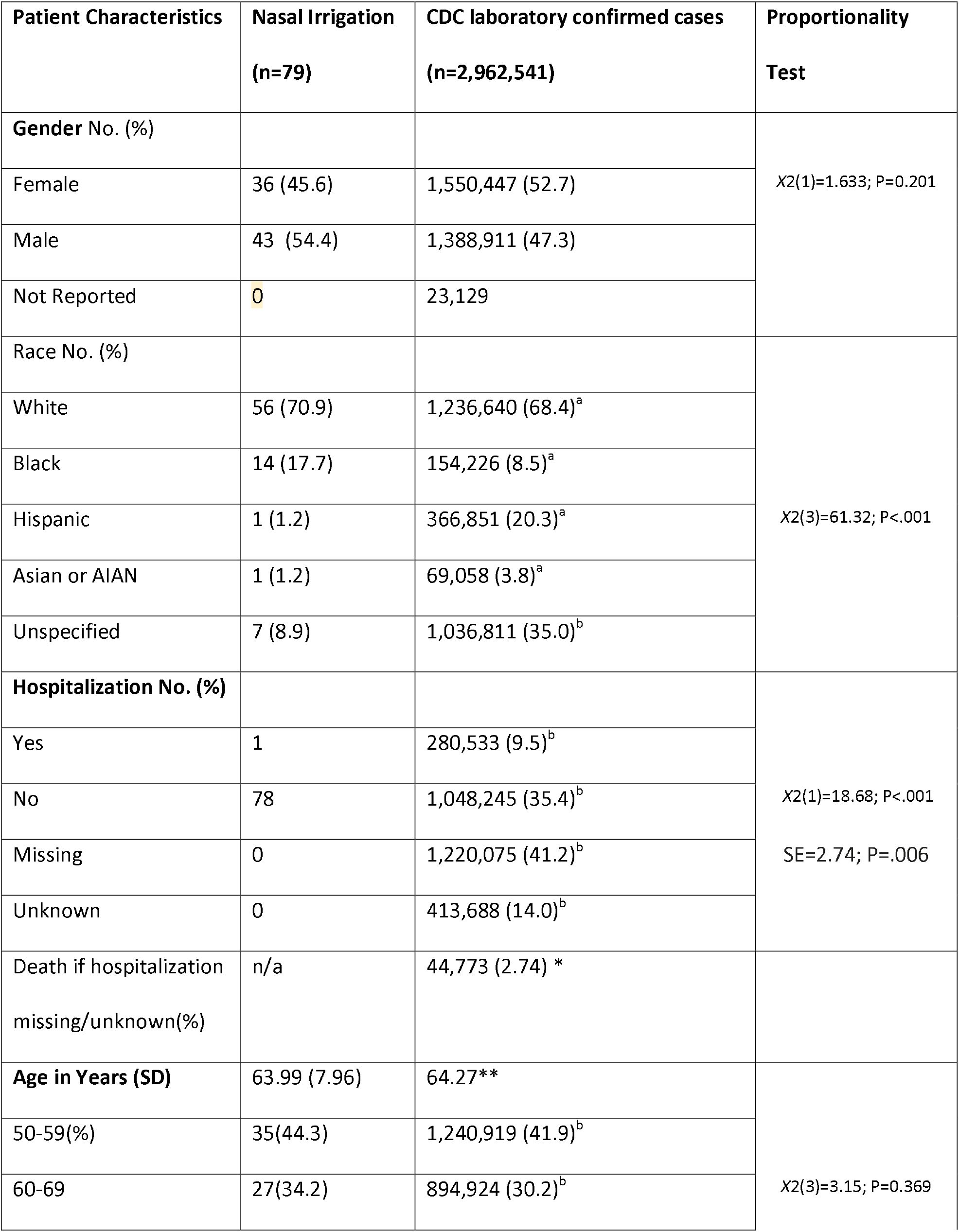
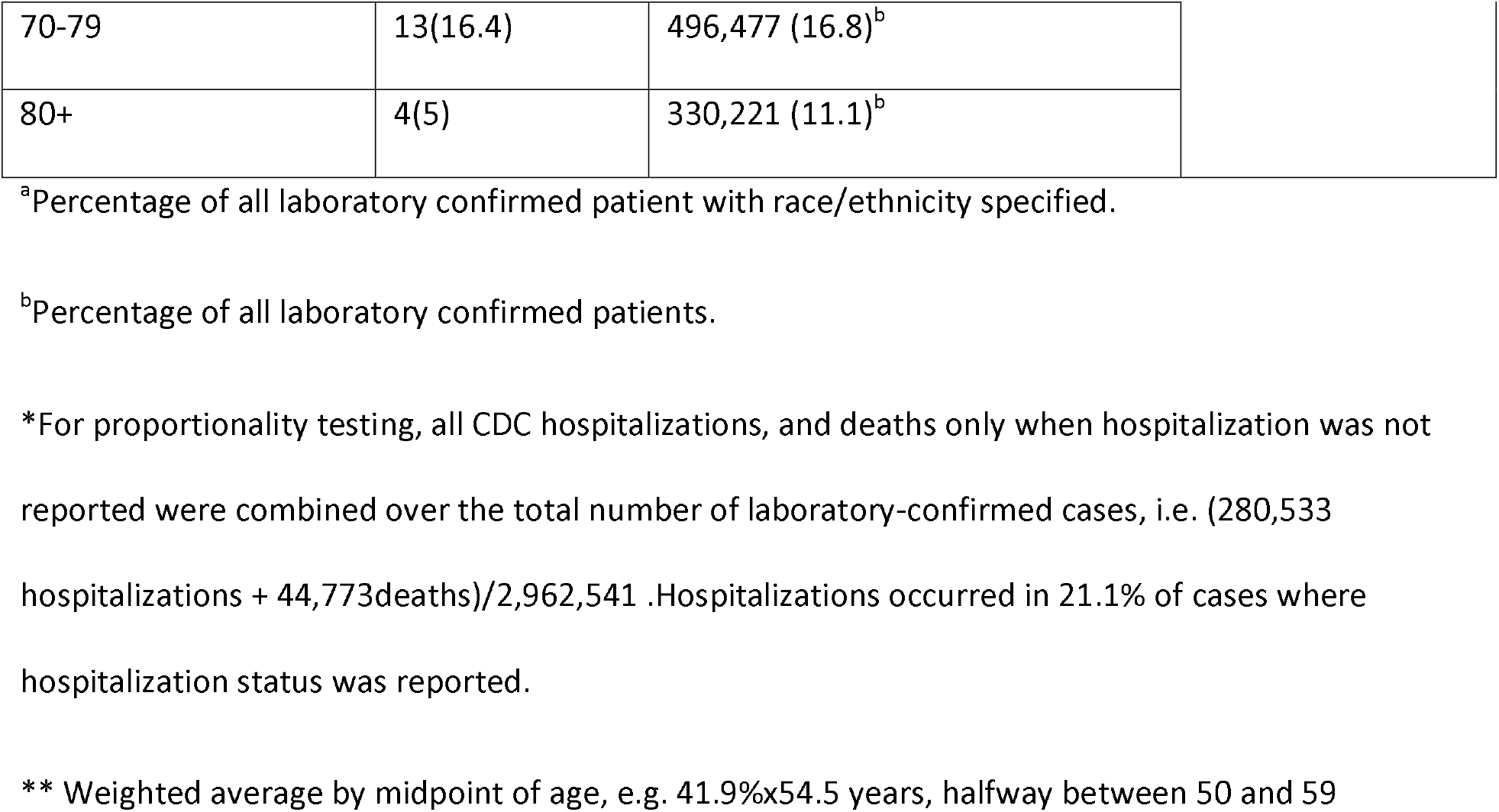

**Table 2:** All participants with completed intake surveys (n=53)

| **Patient Characteristics** | **Nasal Irrigation (n=53)** | **CDC laboratory confirmed cases (n=2,962,541)** |
| --- | --- | --- |
| **Days of illness prior [IQR]** | 4[2,6) | n/a |
| **Age years (SD)** | 63.7 (8.34) | 64.27 |
| **BMI kg/m2 (SD)** | 30.3 (6.75) | n/a |
| **Pre-existing condition No. (%)** | Any (68) | Any(62)<br>Yes: 236701<br>No: 144359<br>Missing data: 2556239 (87%) |
| Obesity by self-report | 11 (20.8) | n/a |
| Obesity by self-report or BMI* | 24 (45) | n/a |
| Hypertension | 23 (43.4) | n/a |
| Asthma | 3 (5.7) | n/a |
| Diabetes | 6 (11.3) | n/a |
| Immunocompromised | 2 (3.8) | n/a |
| None | 17 (32) | n/a |
| Multiple conditions | 22 (41.5) | n/a |

\*Mean BMI 30.3. Median 28.9 [IQR 25.6, 33.1]

Twelve patients received their materials but didn’t record their first irrigation until the following day. Of 631 daily online surveys, patients reported irrigating once per day (7.29%), twice daily (88.43%), or none (4.25%), averaging 1.79 irrigations per day. (Table 3) Patients were asked to take pictures of used irrigation materials to corroborate irrigation, but the number of used packets over time became difficult to assess for confirmation. Five participant provided compliance information at phone calls due to difficulty interfacing online. After enrollment, 11 patients complained of discomfort or spotty epistaxis, with four discontinuing irrigation. (Table 3)

**Table 3.**
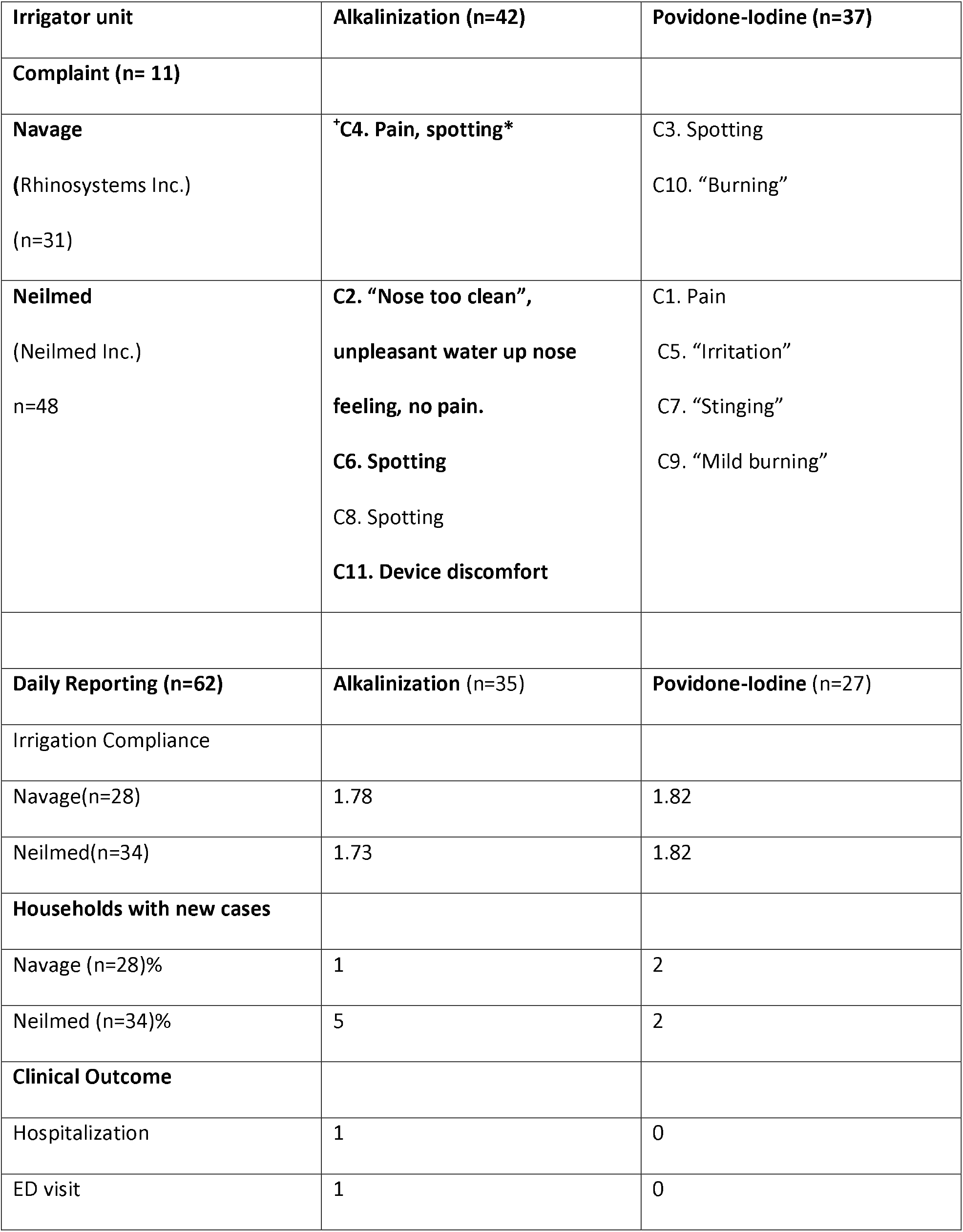
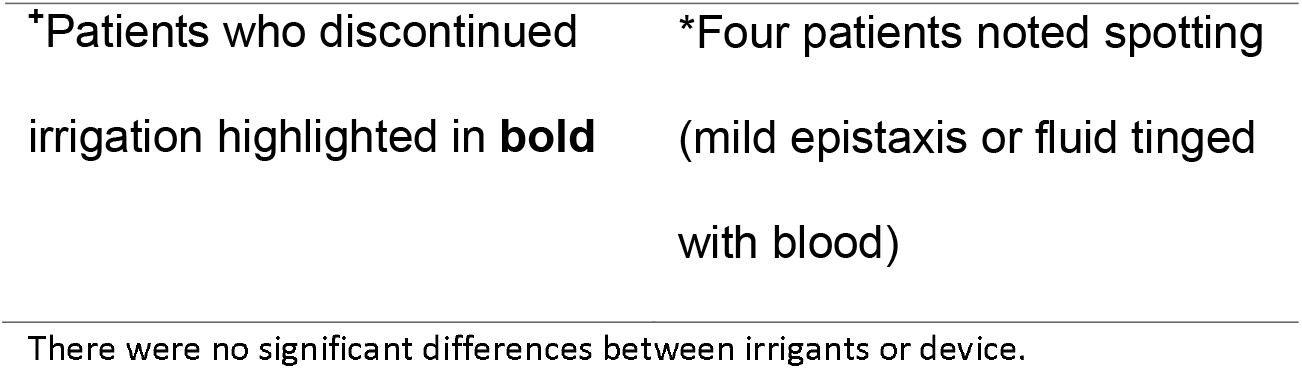
Outcomes by Irrigant and Unit

| **Irrigator unit** | **Alkalinization (n=42)** | **Povidone-Iodine (n=37)** |
| --- | --- | --- |
| **Complaint (n= 11)** |  |  |
| **Navage**<br>(Rhinosystems Inc.)<br>(n=31) | <sup>+</sup>**C4. Pain, spotting\*** | C3. Spotting<br>C10. "Burning" |
| **Neilmed**<br>(Neilmed Inc.)<br>n=48 | **C2. "Nose too clean",**<br>**unpleasant water up nose**<br>**feeling, no pain.**<br>**C6. Spotting**<br>**C8. Spotting**<br>**C11. Device discomfort** | C1. Pain<br>C5. "Irritation"<br>C7. "Stinging"<br>C9. "Mild burning" |
| **Daily Reporting (n=62)** | **Alkalinization (n=35)** | **Povidone-Iodine (n=27)** |
| Irrigation Compliance |  |  |
| Navage(n=28) | 1.78 | 1.82 |
| Neilmed(n=34) | 1.73 | 1.82 |
| **Households with new cases** |  |  |
| Navage (n=28)% | 1 | 2 |
| Neilmed (n=34)% | 5 | 2 |
| **Clinical Outcome** |  |  |
| Hospitalization | 1 | 0 |
| ED visit | 1 | 0 |

|  |  |
| --- | --- |
| *Patients who discontinued irrigation highlighted in **bold** | *Four patients noted spotting (mild epistaxis or fluid tinged with blood) |

There were no significant differences between irrigants or device.

Presenting symptoms present in over 50% of patients included fever, muscle aches, congestion, and headache. There were no statistical differences in symptomatic outcomes by irrigation unit used or irrigant additive, however symptoms resolved for all but 8 patients (12.9%) over the 14 day assessment period.

Ten participants (12.7% by intention to treat) had household contacts who tested positive at least one day after enrollment, compared to 18.8% in a published meta-analysis.^12^ There was no difference in risk of household spread by additive or irrigation unit. (Table 3)

## Discussion

Our results support that pressurized nasal irrigation reduces the likelihood of hospitalization in high-risk COVID-19 + patients, suggesting a safe and over the counter measure with potentially vital public health impact. Nationally, the reduction from 11% to 1.3% as of November 2021 would have corresponded in absolute terms to over 1,000,000 fewer older patients requiring admission. While one study found 47% of those 50 and older had continued symptoms 14 – 21 days after diagnoses,^16,17^ only 13% in our study had symptoms at day 14. If confirmed in other studies, the potential reduction in morbidity and mortality worldwide could be profound.

Clinically COVID-19 differs notably from previous Coronaviridae: children are less impacted; obesity, diabetes, African American race and hypertension are independent risk factors; the relatively pathognomonic symptom of anosmia is present in up to 80% of patients,^18,19^ and the duration from infection to severe symptoms is prolonged.

The clinical differences in presentation reflect SARS-CoV-2’s primarily nasal entry and nasopharyngeal replication. Olfactory neuroepithelium ACE2 is expressed at 700 times the expression in lungs.^19,20^ Conditions increasing nasal ACE2 expression (obesity, hypertension, pollution) or sinus size (age, male sex) correlate with increased severity, further supporting targeting viral fusion in the nasopharynx.^21-25^ In contrast, populations lacking fully developed sinus area (children), with a high baseline practice of nasal irrigation (Laos, Vietnam), or higher mask compliance have decreased severity.^26,27^ Together, the nasal cavity size, ACE2 expression and variolation explanation could account for lower pediatric severity and spread.^28^ The degree of methylation of the ACE2 receptors (and thus stiffness and ease of viral attachment) is related to both race and epigenetic stress.^29,30^ Thus, increased virulence correlating with increased area, quantity and stability of the spike proteins supports the mechanical target hypothesis. Given the local cell-to-cell rather than hematogenous spread and delay in activation of lung TMPRRS2,^31^ mechanically debriding viral particles lodged in the ACE2 receptor, but not yet fused, could reduce severity. Furthermore, the variation in severity with methylation implies that not all particles become securely attached. The size variations in the entire nasal cavity, rather than just anterior nares, support the concept that full nasal cavity irrigation may be superior to nasal spray. Finally, the number of asymptomatic cases and the correlation of illness severity with viral load implied that even after PCR positivity, a window exists wherein lowering the viral load through irrigation could be clinically advantageous.

Ignaz Semmelweis pioneered handwashing to remove bacteria in 1847. In emergency medicine and surgery, debriding infectious material with copious high-powered irrigation is standard practice. While nasal irrigation reduced symptoms of other *Coronaviridae*, flu,^4^ and bacterial carriage in otolaryngology^32,33^, pathology from local spread and aspiration and the continued production of viral load locally suggest a potentially greater impact on COVID-19. Association of viral load with severity ^27,34,35^ suggests a different kind of cumulative pathology related to immune response, as well as the potential for reducing severity after the fact by debridement. Multiple studies have demonstrated immediate viral load reductions in vitro and in vivo with direct oral or nasal application of antivirals,^8-10^ or the theoretical benefit of lavaging and gargling.^36-38^ One study of povidone-iodine gargles and sprays in 24 patients did not show a significant reduction in viral load, but the age difference of 23 years between control and intervention groups calls randomization into question.^39^ An interim analysis of a twice daily nasal irrigation trial in 45 young adults showed initial clinical improvement,^17^ but was ultimately underpowered.

To our knowledge this is the largest prospective clinical trial using both twice daily large volume irrigation with a virucidal arm, and with documented adherence to irrigation. Moreover, the older and higher risk population in this study may be most relevant to reducing morbidity and mortality.

## Limitations

The primary limitations to our study are generalizability and risk of bias in the comparison dataset. Without a matched control group, our sample may differ from the CDC database. Sex and age were not significantly different, but too many patients were missing race/ethnicity and pre-existing conditions to meaningfully evaluate. However, where there were differences our sample was historically more at risk: obesity prevalence exceeded the national average, and over 25% of our patients were Black or did not want to report race, almost double the national average.

The greatest risk of bias comes from preferential reporting of cases with hospitalization or death to the CDC dataset, artificially raising the appearance of severity.^40^ However, our conservatively calculated CDC admission rate of 9.47% reflects the outcomes in other prospective randomized controlled trials: in a younger, thinner group of patients (average age 50, BMI 29), fluvoxamine reduced hospitalization or death from 16% to 11%.^41^ In a healthier cohort evaluating monoclonal antibodies, Chen et al found a 15% admission rate in patients 65+ or with BMI > 35, similar to our population with average age 64 and BMI >30.^42^ Socioeconomic challenges and larger minority populations have higher admission rates. In a similar health system to ours, Price-Haywood et al found a 39.7% admission rate; a Cochran database of minority patients’ admission rates in similar time periods and demographic location to our enrollment period consistently found admission rates as high as 60%.^43,44^

While irrigation could be an effective mechanical protection against variants in vaccinated people, adoption of a new hygiene intervention – or any intervention – is a barrier. Of the 537 patients contacted, 28 (5.2%) did not want to perform nasal irrigation. Of those who initiated irrigation, most continued twice daily use, but eleven had concerns about irrigating that were communicated to our staff. While only four discontinued irrigation, without the frequent calls and coaching adherence in the general population could be lower. The need for boiled, distilled or filtered water could also be ignored, introducing a new risk.

Our study was underpowered to detect improvement by additive. While low povidone-iodine concentrations are safe up to five months,^7^ studies using tenfold higher concentrations for gargling noted transient thyroid stimulating hormone changes.^39^ For prolonged use, thyroid function testing may be warranted. Studies of alkalinity have not supported efficacy in reducing viral fusion, however recent evaluation of the Omicron variant suggest hypertonic saline may add benefit to irrigation.^3^

## Conclusion

As an intervention, pressurized nasal irrigation showed promise to reduce the severity of COVID-19 infection in high-risk patients when initiated within 24 hours of a positive test. As large unvaccinated populations pressure evolution of variants, an effective mechanical outpatient intervention to reduce viral entry and hospitalizations can save lives and reduce the stress on hospital staff. Further research into the frequency and adjuvants of irrigation will be important not just for this pandemic, but for future viruses to come.

## Supporting information

Statistical Analysis Plan and Protocol

## Data Availability

Data is available from the University of Augusta Department of Emergency Medicine Research Office, and online from the CDC COVID-19 Case Surveillance Public Use Data

https://data.cdc.gov/Case-Surveillance/COVID-19-Case-Surveillance-Public-Use-Data/vbim-akqf/data

## Notes

### Competing Interest Statement

The authors have declared no competing interest.

### Clinical Trial

NCT04559035

### Funding Statement

Materials were provided by Neilmed Inc. and Rhinosystems Inc. The investigator-initiated study was supported by funding from the Bernard and Anne Gray Donor Advised Fund Community Foundation for Greater Atlanta, Neilmed Inc., and Rhinosystems. No authors received payment or services from a third party for any aspect of the submitted work.

### Author Declarations

Institutional Review Board Office Augusta University 1120 15th St., CJ-2103 Augusta GA 30912-7621 Phone: 706-721-3110 http://www.augusta.edu/research/irboffice/

### Summary of Updates

Updated data from the CDC Dataset improved reported hospitalization information (accessed 11/4/2021); verified all irrigation patients hospitalization status; corrected one hospitalization attributed in alkalinization group to betadine.

