## Supplementary material for "Rapid initiation of nasal saline irrigation to reduce severity in high-risk COVID+ outpatients: a randomized clinical trial compared to a national dataset observational arm": Statistical Analysis Plan and Protocol

eAppendix. Supplemental Methods

This supplement contains the following items:

1. Final statistical analysis plan, summary of protocol changes and approvals.
2. Recruitment detail
3. Final Protocol

**Rapid Initiation of Nasal Saline Irrigation to Reduce COVID-19 Severity:**

A single center randomized controlled non-blinded clinical efficacy study

evaluating 0.1% Povidone-Iodine compared to alkalinized nasal irrigation BID

for the reduction of COVID-19 severity in COVID+ outpatients.

Principal Investigators: Amy L. Baxter

Co-Investigators: Kyle Schwartz, Matthew Lyon, Richard Schwartz

Department of Emergency Medicine Medical College of Georgia at Augusta University

**CONTENTS**

1.	Administrative	3

2. Introduction	3

1.1 Background and Rationale	3

1.2 Objectives	3

2.	Study Methods	4

2.1	Trial Design	4

2.2	Randomization	4

2.3	Sample Size	4

2.4	Statistical Framework	5

2.5	Statistical Interim Analyses and Stopping Guidance	5

2.6	Timing of the Final Analysis	5

3.	Statistical Principles	5

3.1	Confidence Intervals and P-Values	5

3.2	Protocol Deviations	5

4. Trial Population	5

4.1	Screening Data	5

4.2	Eligibility	5

4.3	Recruitment	6

4.4	Withdrawal/Follow-up	6

4.5	Baseline Patient Characteristics	6

5.	Analysis	6

5.1	Outcome Definitions	6

5.2	Analysis Methods	7

5.3	Missing Data	7

5.4	Additional Analyses	7

5.5	Harms	8

5.6	Statistical Software	8

11.	Appendix 1 Full Recruitment Demographics and Protocol	9

12.	Appendix 2 Final Protocol	11

**Rapid Initiation of Nasal Saline Irrigation to reduce COVID-19 morbidity and mortality (RINSI) – Statistical Analysis Plan**

1. **Administrative Information**
   a. **Trial Registration Clinical Trials.gov** NCT04559035
   b. **Funding: Investigator initiated**
   c. **Statistical Analysis version 9.7.21-2 Protocol Version 1.14.21-3**
   d. **SAP Revision history**

**Protocol Version 7-27-20 –** Prospective randomized controlled trial of alkalinization compared to povidone-iodine twice daily nasal irrigation initiated on the same day of notification of positive COVID-19 Test to reduce morbidity and mortality.

**Protocol Version 10-16-2020 –** Change to allow healthcare workers to participate. Due to disproportionate difficulty enrolling Black population, protocol changes allowing enrollers to leave a message and re-call, revising study flyer to emphasize benefit and moving “clinical research” language lower on the page.

**Interim Review of Primary Outcomes 12-6-2020 –** Due to introduction of monoclonal antibodies, anticipated vaccination, and staffing, decision to do interim analysis early. Discovery that zero patients had been hospitalized, compared to an expected 25% and a rate of 16.4% in the state. Recalcluation of power analysis grouping all nasal irrigation patients compared to matched controls.

**Protocol version 1-11-2021 –** Change to case-control analysis of primary outcome comparison of hospitalization and death to matched unenrolled controls using 5-hospital EHR data. IRB approved 1.14.2021.

**Analysis change 8-7-2021** – Change to primary outcome analysis as a case-cohort of risk probabilities. After contractual barriers to accessing EHR, decision to use national CDC dataset for binomial analysis of probabilities between original group and laboratory confirmed cases. Final analysis based on dataset accessed 8-20-2021

e. **Contributors**
   i. **Statistician responsible:** Kevin M. Swartout Department of Psychology, Georgia State University, Atlanta GA
   ii. **Principal Investigator and SAP author:** Amy Baxter, Department of Emergency Medicine, Augusta University, Augusta GA USA

2. **Introduction**
   a. **Background and rationale** Research suggests that acidic pH and nasal viral load contribute to replication and disease severity of COVID-19. We plan to test saline nasal irrigation of 240cc with either alkalinized nasal saline or 0.1% virucidal povidone-iodine twice daily in participants recently testing positive for SARS-CoV-2. We hypothesize that nasal irrigation debridement with either the basic or virucidal intervention will be correlated with reduced COVID-19 progression to hospitalization, and reduction in symptom severity, supportive oxygen, and death in COVID-19 positive patients 55 years of age and older.
   b. **Objectives-** To determine if twice-daily saline nasal irrigations with either 0.1% povidone-iodine or 2.5 mL of baking soda in one of two over-the-counter irrigation devices in patients tested positive for COVID-19 improves clinical outcomes:
      i. **Primary Outcome:** To assess reduction in COVID-19-related hospitalizations and death compared to a national dataset.
      ii. **Secondary Outcomes:**
          1. Assess reduction in hospital visits, symptoms, admission, supplemental oxygen use, ICU admission, and death in patients with alkalinization or virucidal additives to nasal irrigation.
          2. Evaluate impact of nasal irrigation compared to published historical reports and meta-analyses of outcomes in similar patient groups including household spread.
          3. Evaluate impact of race, days of symptoms, and pre-existing conditions on utility of nasal irrigation to reduce adverse outcomes of any kind.
3. **Study Methods**
   a. **Trial design -** a consecutive prospective, randomized parallel group trial to evaluate the efficacy of either alkalinization or povidone-iodine twice daily nasal irrigation initiated on the day of positive test notification for reduction of COVID-19 symptoms and disease progression to hospitalization or death**.** The enrolled participants were also compared to a national dataset as a nested case-cohort.
   b. **Randomization -** Block randomization to the twice daily irrigation with alkalinization or povidone-iodine was done by random number generator (random.org) stratified by sex. With odd numbers signifying alkaline and even povidone-iodine, numbered opaque envelopes were prepared in separate sequences for male or female participants to be opened after consent, indicating the appropriate additive to be given to the patient. The list of positives for the day from the lab were called in sequential order to obtain a consecutive sample that met all inclusion criteria and did not meet any exclusion criteria. On days with potential participant volume exceeding 10, the call order was randomized by random number generator.
   c. **Sample Size -** We assumed a baseline admission rate of 25%, with a small to moderate impact of the additive to the irrigant (Cohen’s d=3). To reduce hospitalizations to 10%, 100 patients per group would be required for 80% power. Initial assumptions by the statistician were that irrigation additive as well as mechanical unit would be variables, thus the IRB approved protocol was for 400 patients. Published support that any pressurized unit was sufficient reduced the target enrollment to 200.

After determining a zero hospitalization rate in early December, the trial’s revised target enrollment was 79 irrigation participants with 158 matched controls. Assuming that 25% of the matched control participants and 10% of the treatment participants would visit the hospital (60% reduction), 79 participants in the nasal irrigation condition would have .80 (80%) power to detect that proportional difference. Power was calculated two ways, by running a simulation comparing the likelihood of hospitalization, and using a z-test comparing independent proportions. Both analyses assumed reduced variation in proportions due to accounting for the correlates of testing time, age, sex, and race, and resulted in similar calculations. The inability to evaluate and match underlying conditions in the control group could increase variability, thus a 1:2 matching schema was chosen.

d. **Framework -** The RINSI trial was completed using a probability of proportions framework, using an exact binomial test to assess the probability of observing the proportion of patients with morbidity or mortality in this study compared to a national rate. Initial comparisons between groups were conducted to evaluate superiority of either irrigation regimen.
e. **Statistical Interim Analyses and Stopping Guidance -** An interim analysis was anticipated when half of enrollment had been achieved. The availability of monoclonal antibodies changing risk catetories, vaccinations, and end of year staffing concerns led to a decision to do an early interim analysis. With the discovery that none of the patients had been hospitalized, rather than conduct a full analysis a new power analysis was conducted, leading to the decision to enroll up to a target of 79 patients with 1:2 matched controls to speed publishing results.
f. **Timing of Final Analysis -** The final analysis began after the 28-day outcome was completed for all participants January 18th. During the blinded collection of outcome data from the five hospitals in the catchment area, the discovery of blocked access due to legal agreements was made. Several months of discussion and meetings were not able to resolve access to the matched control data. In May and June, discussions with the Georgia Department of Public Health were undertaken to get access to case and hospitalization data without results. August 7th a CDC correspondence directed the team to the national database as a comparison cohort stratified by age, allowing the final analysis comparing the outcomes in enrolled patients compared to the CDC dataset last accessed 8/20/2021.
g. **Time points at which outcomes were measured:** Primary outcomes were measured 28 days after enrollment. Secondary outcomes of symptoms were collected daily via online survey for the first 14 days. Subjects were called on Day 2, 7, 14, and 28 to check on current symptoms and outcomes.

4. **Statistical Principles**
   a. **Confidence Intervals and P Values:** alpha will be 0.05 and 95% confidence intervals will be reported for relative risk. Standard Error will be reported for exact binomial tests.
   b. **Adherence and protocol deviations:** Symptom outcomes will be reported for patients completing the daily symptom diary. Adherence to irrigation was recorded in daily online surveys, and confirmed with uploaded photographs of used saline packets or pods, or phone calls for patients unable to upload photographs.
   c. **Analysis populations:** The primary outcomes of hospitalization and death were by intention to treat. Adherence, symptom, and household spread will be reported for patients filling out a minimum of two daily surveys, and all completing initial survey.
5. **Trial Population**
   a. **Screening Data:** We will report the number of patients with positive tests, the number reached, the number screened and reasons for non-enrollment.
   b. **Eligibility:**
      i. **Inclusion Criteria** - Capable of understanding and providing informed consent using remote consent; Willingness and physical capacity to initiate nasal irrigation and to adhere to the protocol; Willing to give additional contact phone number of another person who will know the health status of the participant and agree to be contacted if needed for follow-up; 55 years of age or older; Has access to and the willingness and ability to adhere to the technological requirement of the study i.e. able to use a smartphone for voice and text and email and access to the internet at home; English speaking; Positive rapid COVID-19 test performed the day of or the day before enrollment
      ii. **Exclusion Criteria** - Currently doing daily nasal irrigation; Current supplemental oxygen therapy; Unwillingness to try nasal irrigation or use nasal irrigation twice a day; Nasal surgery within the past year or chronic sinusitis; Prior COVID-19 infection or positive test >1 day before present; Symptoms longer than 7 days prior to testing as reported to researchers; Allergy to iodine or shellfish; Participation in another prospective COVID related research project (clinical trial).
   c. **Recruitment:** Information to be included in the CONSORT flow diagram: See Figure 1. Recruitment procedures are in Appendix 1. Reasons for refusal are in Appendix 2.
   d. **Withdrawal and follow-up:** Those discontinuing nasal irrigation will be reported with reasons.
   e. **Baseline Characteristics:** characteristics of all irrigation patients compared to CDC database characteristics will be reported, including sex, age, race, pre-existing conditions with mean and standard deviation where possible.
6. **Analysis**
   a. **Outcome Definitions** –
      i. **Primary Outcome**: hospitalization or death from COVID-19 within 28 days of enrollment – Obtained by self-report, report of family member, and confirmed by chart review. For the purposes of the study, COVID-19 related chief complaints will include any of the top five symptoms associated with emergency department presentation: Cough, Fever (subjective or >100.4°F/38°C), Myalgia, Headache, or Dyspnea. Other ED presentations resulting in discharge without subsequent admission will not meet the criteria of morbidity severity related to this study. In cases where patients are admitted, discharge diagnosis or cause or contributing cause of death will be considered for categorization of COVID-related or not-COVID related. In cases where potential for COVID-19 related admission or death is ambiguous, a member of the research team (MD) blinded to group will be asked to assess the information gathered from the chart and make a determination of COVID or non-COVID related.
      ii. Secondary Outcomes – Obtained by online surveys, in addition to confirmatory phone call information collected day 2,7,14, and 28.
          1. Duration of COVID-19 symptoms (loss of smell, fatigue, presence or absence of fever >100.4, subjective fever, chills, muscle aches, runny nose, cough (new onset or worsening of chronic cough), shortness of breath, nausea or vomiting, headache, loss of smell, abdominal pain, diarrhea (>3 loose/looser than normal stools/24 hours)
          2. Confirmed or symptomatic household members with new cases after initiation of daily surveys
          3. COVID-19 related ED visits, supplemental oxygen use, ICU admission
          4. Dose-response of daily irrigation related to symptoms
          5. Adverse events related to irrigation, including discomfort not leading to or leading to discontinuation and reason
   b. **Analysis methods** –
      i. **Primary Outcome** -To compare observed hospital admission rates among participants compared with national rates of severe disease (admission or death) published by CDC, the binomial test was used. As it is an exact test, Clopper-Pearson confidence intervals were used giving a SE and P value. Relative risks were calculated for bad outcomes compared to the total number of cases, and hospitalization only when status was reported to give lower and upper ranges of impact given our concerns of reporting bias in the dataset. Chi squares were calculated for differences between demographics in the enrolled group and the national dataset.
      ii. **Differences by irrigant method.** Prior to combining the randomized groups to compare with the national dataset, the plan was as follows: to examine sex and racial differences on the dependent variables, *t*-tests will be used unless non-normal distribution is noted, in which case Mann Whitney U will be used. Baseline measures of duration, pre-existing conditions, and symptoms will be compared across the conditions to ensure that the randomization process was effective. Analyses of compliance by device and the previously mentioned dependent variables will be conducted to determine whether the brand of device impacted outcomes. If any of these analyses result in significant findings, they will be considered as covariates in subsequent analyses. To test the effect of treatment group in disease severity, Kaplan-Meier survival plots will be calculated. Pearson’s correlation coefficient will assess if an inverse linear relationship exists between number of irrigation uses and illness severity outcomes.
   c. **Missing Data** – Only laboratory confirmed cases were used. Hospitalization status yes or no was reported for 43% of laboratory-confirmed CDC case records in correlating age ranges and the same enrollment time period. To avoid overestimating by reporting bias, we calculated the admission rate retaining all laboratory-confirmed cases in the denominator, including when hospitalization status was missing or unreported.

(Hospitalizations reported yes)

______________________________________________________

(Hospitalization Yes + No + Unknown + Missing)

Because there were no deaths in our sample, we could not calculate a comparative rate with the national database. To capture the burden of mortality which likely was or should have been preceded by hospitalization, we included deaths as an additional measure of severity only when hospitalization was unknown or missing. Thus, the rate of deaths is under-estimated.

Deaths reported

___________________________________

(Hospitalization Unknown + Hospitalization Missing)

We did not include deaths in cases where hospitalizations are reported in our analysis to avoid counting outcomes with increased severity twice. Thus the comparison rate used for binomial testing was:

Hospitalizations + Death when hospitalization unknown or missing

(Hospitalizations Yes + No + Unknown + Missing)

d. **Harms –** Adverse events are summarized in Table 3**.**
e. **Statistical Software -** IBM SPSS Statistics for Windows, Version 27.0. Armonk, NY: IBM Corp; (MedCalc Software Ltd. https://www.medcalc.org/calc/relative_risk.php (Version 20.009))

Appendix 1: Final Full Recruitment Methods and Demographics

Recruitment

There will be two groups of participants: an active (exposed to intervention) group and a passive control group. The passive “nonexposed” control group (not interested in enrolling or not able to be contacted ) will be people 55 years of age and older who tested positive for COVID-19 during the time period of the study that were not enrolled in the study. The names and birthdays of these individuals will be collected so that approximately 4 weeks later the EMR can be searched to see if they sought care at an Emergency Department or were hospitalized for COVID-19 related symptoms. The other group of participants will be the active group.

Active participants will be community outpatients who have tested positive for COVID-19. Participants will be recruited by obtaining a list of outpatient positive results from the AU testing center of patients age 55 years or older. Persons on this list will be called by a member of the research team and asked if they are interested in participating in the study. If they do not pick up the phone a voice mail will be left (see script) and they will be called back up to two additional times (unless the participant calls us back first).

At the AU local testing site potential participants will be given an informational flyer about the study. The flyer will also be made into posters that will be posted at the testing site. The flyer will explain that if they test positive they may be eligible to participate in a research study evaluating the effects of nasal irrigation on symptoms associated with COVID-19, and instructing them to call the number on the flyer for more information when their positive test result returns. The flyer will also provide information about the on-line health care portal which can be used to obtain test results more quickly. In addition to the flyer a banner will be added to the health care portal informing users about the study and providing contact information where they can learn more. A minimum of once a day a member of the research team will obtain the list of positive results from the on-site testing facility. Those individuals who tested positive and who meet study inclusion criteria will be called and asked if they are interested in participating in the study (see phone script). If patient is interested in hearing more the study will be explained and remote consent will be obtained. Participants will be asked to enter the phone number of the research team into their cell phone in order easily identify future calls.

4.1.1. Eligible participants will be consented remotely and their study materials (consisting of a nasal irrigation device (Neilmed sinus irrigation bottle or Navage unit) with 28+ accompanying saline pods/packets, two gallon jugs of distilled water, a physical copy of the consent form, an instructional sheet, and the randomly allocated additive (baking soda or betadine) with a ½ tsp scoop), will be delivered to their residence by a member of the research team using COVID-19 precautions (masks, maintaining 6 ft. or more physical distance, door drop off) later that day.

The Neilmed and Navage irrigation devices will be distributed on alternating days with the goal of distributing them evenly among participants to avoid conflicts of interest. Neilmed will be distributed on Monday, Wednesday, Friday, and Sunday, while Navage will be Tuesday, Thursday, and Saturday.

After having the study procedures explained to them and consenting to the research, the Active participants will be provided with an instructional sheet along with their research materials. This sheet will contain detailed instructions on how to mix ½ tsp of their allocated additive into the saline solution and perform a nasal irrigation, and include instructions on how to view a Youtube video demonstrating how to conduct a nasal irrigation with the provided device. The Active participants will be instructed to perform nasal irrigation twice a day for the next 14 days, ideally with a window of at least six hours between irrigations. The participants will be provided with instructions on how to access the online surveys, which are distributed to them each day through their email.

Irrigation data will be collected over Qualtrics surveys, which will be delivered on a daily basis to the participants email address for a period of 14 days following enrollment. In addition to the number of irrigations performed each day, the Qualtrics surveys collect background information related to demographic, medical history, current medications, current symptoms, and daily adherence by way of a photograph of the used saline containers. The participants will be called for follow-up on days 2, 7, 14, & 28. Participants who have not filled out a survey for two consecutive days will be called at any time during the first 14 days, and the total number of irrigations performed will also collected verbally over the phone on the 14th day of participation if the patients are unable to upload pictures to the database. On the 28th day, the participants or their designated follow-up person will be asked about hospitalizion or medical care related to COVID-19 within the study period. A chart review of the EHR containing data from the five community hospitals will also be conducted to determine the same.

Appendix B – Final Protocol

**Nasal Irrigation to Reduce COVID-19 Morbidity**

NCT04559035

Revision: 1/11/2021

**Protocol Title:** [COVID-19] Saline Nasal Irrigation to Reduce COVID-19 Severity

**Principal Investigators:** Amy Baxter, MD; Matt Lyon, MD

1. Objectives

Describe the purpose, specific aims, and hypothesis:

Research suggests that acidic pH and nasal viral load contribute to replication and disease severity of COVID19. We plan to test saline nasal irrigation with either Alkalinized nasal saline or ½ tsp of the virucidal povidone-iodine twice daily in patients recently testing positive for SARS-CoV-2. We hypothesize that nasal irrigation debridement with either the basic or virucidal intervention will be correlated with reduced COVID-19 progression to hospitalization, and reduction in symptom severity, supportive oxygen, and death in COVID-19 positive patients 55 years of age and older

**Specific Aims:**

Evaluate the number of twice-daily saline nasal irrigations with either povidone-iodine or ½ tsp of baking soda in one of two over-the-counter irrigation devices in patients tested positive for COVID-19 with regards to clinical outcomes.

Technical Objective 1: To assess reduction in COVID19-related hospital visits, symptoms, admission, supplemental oxygen use, ICU admission, and death in patients with differing nasal irrigation frequency.

Technical Objective 2: Assess reduction in hospital visits, symptoms, admission, supplemental oxygen use, ICU admission, and death in patients with alkalinization or virucidal additives to nasal irrigation.

- Evaluate impact of nasal irrigation compared to published historical reports and meta-analyses of outcomes in similar patient groups including household spread.
- Evaluate impact of nasal irrigation on COVID-19 related hospitalization compared to hospitalization data in matched control group.
- Evaluate dose-response of irrigation to clinical outcomes as noted above.
- Evaluate impact of race, days of symptoms, and pre-existing conditions on utility of nasal irrigation to reduce adverse outcomes of any kind.

Click here to enter text.

*Describe the background and rationale for the study:*

The severe acute respiratory syndrome coronavirus 2 (SARS-CoV-2) pathogen is a single stranded positive sense RNA virus, with a similar spike receptor binding mechanism as SARS-CoV and MERS-CoV. Clinical coronavirus disease 2019 (COVID-19) caused by the pathogen differs in several notable ways from previous viral coronaviridae: younger patients are dramatically less impacted than the elderly; obesity, diabetes, race and hypertension are independent risk factors; male patients fare more poorly than females; the duration from infection to mild symptoms to severe symptoms averages 14-17 days; anosmia is present in up to 80% of patients; and asymptomatic transmission is common.

Recent work demonstrates that the attack entry overwhelmingly occurs via respiratory or aerosolized droplets entering in the nasal mucosa at the ACE2 receptors in ciliated epithelial cells,(1) explaining many of the clinical pathogenic differences. Nasal ACE2 receptors are upregulated in obesity, diabetes and hypertension. Children have minimally developed sinuses, lower expression of ACE2 and TMPSSR genes, and much smaller nasal cavities compared to adults, with males over 70 having significantly larger cavities and mucosa than women.(2) Furthermore, viremia is infrequent, supporting the hypothesis of local mechanical spread of the virus toward the lungs as a predominant means of infectivity. Alternately, genetic expression of the ciliated nasal ACE2 and TPMSSR genes has been shown to activate expression in the lungs.

The preponderance of nasal entry and the slow time frame for sufficient viral replication suggest that a topical debridement may reduce the viral load and ability of the virus to replicate sufficiently, allowing time for immune response without inciting an overwhelming cytokine storm. While drug treatment tests are ongoing, the nasal entry point and previous work in reduction of viral load and symptoms with other pathogens suggest topical antiviral and pH changing interventions could be uniquely effective at reducing attachment, viral fusion, and viral load, thus reducing morbidity and mortality.

Significance

If this study yields positive results and shows nasal irrigation to be a safe, inexpensive and accepted method for reducing COVID19 severity in a racially diverse and economically challenged population at higher risk for morbidity and mortality, it will potentially save hundreds of thousands of lives.

Research & Development by others

1) Nasal viral load is highly proportional to infectivity.(4) Mechanically flushing out SARS-CoV-2 would be expected to reduce the viral load.

2) The nasal mucosa is specifically the site of SARS-CoV-2 entry; ACE2 receptors are highly expressed in nasal epithelial cells.(1) Indeed, an intranasal inhibitor of spike fusion eliminated disease in mice exposed to SARS-CoV-2. “Intranasal application of EK1C4 before or after challenge with HCoV-OC43 protected mice from infection, suggesting that EK1C4 could be used for prevention and treatment of infection by the currently circulating SARS-CoV-2 and other emerging SARSr-CoVs.”(3)

3) Mechanical reduction of cellular infection: The viral lipid layers of SARS-CoV-2 need to fuse after the spike attaches to ACE2 receptors, so physical flow of nasal irrigation may reduce likelihood of intracellular fusion and thus infection. Per the Nature article, “Our data highlight the possibility that viral transmissibility is dependent on the spatial distribution of receptor accessibility along the respiratory tract.”

4) Buying immunologic time: given the apparent mechanical rather than blood-borne slow advance of COVID-2 illness from nasopharnyx to oropharynx to trachea to lung, the possibility exists that reducing viral load through debriding could aid effective immune response in the same way that debriding burns reduces time of healing. People with obstructive illnesses and patients over 80 have dramatically increased lung connections, the collateral ventilation through channels of Lambert and Pores of Kohn. Again, children under 5 do not have this system, lending credence to the surface area/mechanical model of viral severity. The respiratory spread of SARS-COV also spread mechanically, but more efficiently in lungs,(5) supporting interventions prior to respiratory progression.

5) Irrigation works for colds: Nasal lavage (rinsing) seems better than spray. Multiple RCTs show statistically significant results, and the mechanical concepts of COVID lend greater rationale than upper respiratory infections with shorter incubation periods. In addition, nasal irrigation significantly reduce not only load but duration and severity of other viral illnesses even when symptoms had been established as much as 48 hours.(6, 7) Given the slow progression of COVID-19, earlier initiation of irrigation in positive patients not yet requiring hospitalization could have a more marked impact on reducing morbidity.

6) Nasal antimicrobial reduces viral infections in bovine animals: from 28% in those pre-treated with one intranasal NO dose compared to 78% of animals treated with saline placebo.(8) For humans, dilute povidone-iodine is widely used for chronic sinus infections and decolonizing before surgery.(9-12)

7) SARS-CoV-2 may have difficulty fusing with alkaline pH. Previous coronaviridae have needed acidic or neutral pH for the lipid layer between virus and cell to fuse. Alkalinizing the nares may help. The fact that African Americans have more acidic nasal mucosa (6.4 compared to 6.8 in whites) could contribute to the differential infection rate,(13) rendering this intervention helpful in a population with higher attack rates and severity. In addition, drug targets such as zinc, quercetin, and ivermectin act by prohibiting the acidification of the endosomes where viral replication takes place. Providing weak base through mild alkalinization to 7.3 could directly reduce the ability of viral replication.

8) SARS-CoV-2 is sensitive to povidone-iodine solution,(14) as are other viral pathogens,(15) and it is safe for the nasal passages.(9)

9) Mortality in countries where nasal irrigation is widespread is dramatically lower than in countries where irrigation is not standard. Thailand, Vietnam, Laos and Cambodia have populations in which up to 80% of the population commonly use nasal irrigation.(6, 16, 17) Death rates there are fractional compared to other countries which only wear masks or bow in greeting but do not have a cultural habit of irrigation.

FDA regulations. The components of the device – an intranasal irrigation device hand-powered- is rated Class I 510(K) Exempt 874.5550 Product Code KMA. Povidone-Iodine (Betadine) is an FDA approved over the counter drug at strengths of 10% and above and is labeled and indicated for nasal use NDA 019476.

Click here to enter text.

2. Background
3. Inclusion and Exclusion Criteria

*List the inclusion/exclusion criteria:*

*Inclusion Criteria*

- Capable of understanding and providing informed consent using remote consent
- Willingness and physical capacity to initiate nasal irrigation and to adhere to the protocol
- Willing to give additional contact phone number of another person who will know the health status of the participant and agree to be contacted if needed for follow-up
- 55 years of age or older
- Has access to and the willingness and ability to adhere to the technological requirement of the study i.e. able to use a smartphone for voice and text and email and access to the internet at home
- English speaking
- Positive rapid COVID-19 test performed the day of or the day before enrollment

*Exclusion Criteria*

- Currently doing daily nasal irrigation
- Current supplemental oxygen therapy
- Unwillingness to try nasal irrigation or use nasal irrigation twice a day
- Nasal surgery within the past year or chronic sinusitis
- Prior COVID-19 infection or positive test >1 day before present
- Symptoms longer than 7 days prior to testing as reported to researchers
- Allergy to iodine or shellfish
- Participation in another prospective COVID related research project (clinical trial).

Click here to enter text.

4. Number of Subjects/Records/Samples Collected

*Indicate the total number of subjects to be accrued/records reviewed/samples collected across all sites:*

400

5. Recruitment Methods

Describe when, where, and how potential subjects will be recruited:

There will be two groups of participants: an active (exposed to intervention) group and a passive control group. The passive “nonexposed” control group (not interested in enrolling or not able to be contacted ) will be people 55 years of age and older who tested positive for COVID-19 during the time period of the study that were not enrolled in the study. The names and birthdays of these individuals will be collected so that approximately 4 weeks later the EMR can be searched to see if they sought care at an Emergency Department or were hospitalized for COVID-19 related symptoms. The other group of participants will be the active group.

Active participants will be community outpatients who have tested positive for COVID-19. Participants will be recruited by obtaining a list of outpatient positive results from the AU testing center of patients age 55 years or older (See data collection form.) Persons on this list will be called by a member of the research team and asked if they are interested in participating in the study. If they do not pick up the phone a voice mail will be left (see script) and they will be called back up to two additional times (unless the participant calls us back first).

At the AU local testing site potential participants will be given an informational flyer about the study (see recruitment flyer). The flyer will also be made into posters that will be posted at the testing site. The flyer will explain that if they test positive they may be eligible to participate in a research study evaluating the effects of nasal irrigation on symptoms associated with COVID-19, and instructing them to call the number on the flyer for more information when their positive test result returns. The flyer will also provide information about the on-line health care portal which can be used to obtain test results more quickly. In addition to the flyer a banner will be added to the health care portal informing users about the study and providing contact information where they can learn more. A minimum of once a day a member of the research team will obtain the list of positive results from the on-site testing facility. Those individuals who tested positive and who meet study inclusion criteria will be called and asked if they are interested in participating in the study (see phone script). If patient is interested in hearing more the study will be explained and remote consent will be obtained. Participants will be asked to enter the phone number of the research team into their cell phone in order easily identify future calls.

Figure *Flow Chart of Study Recruitment and randomization*

Testing Patient Pool (*n* = 600/day)

Study Sample positive/day 10%+ n=60

Study Sample positive over 55 years (n=24/day)

Measures: Demographic, symptoms, treatments

Block Randomized by sex and race to intervention

*Post Procedure Measures* (*n* = 200): Treatment diary; clinical outcomes called weekly

6. Multiple Site	☐ X N/A

*A site is defined as an institution/organization/university that is collaborating with Augusta University*

*If this research involves multiple sites, specify which is the lead site and describe the roles of each site in the study.*

*If Augusta University will serve as the lead site, indicate that all required approvals are already obtained or will be obtained at each site prior to project implementation. In addition:*

*Describe the processes you have in place to ensure successful coordination of activities among sites. For example, do all sites have the most current version of the protocol, consent document, and HIPAA authorization? How will modifications be communicated to sites and approved prior to implementation? How will participating sites be kept abreast of any problems, interim results, or the eventual closure of the study?*

*Describe the mechanisms you have in place to ensure that all local site investigators conduct the study appropriately and that engaged participating sites safeguard data as required by local information security policies. Please confirm that all non-compliance and /or unanticipated problems associated with the study protocol or applicable requirements will be reported in accordance with local policy.*

Click here to enter text.

7. Reliance Agreements/Single IRB	☐ X N/A

*Reliance agreements (i.e. IRB Authorization Agreement (IAA), Individual Investigator Agreement (IIA), etc.) are formal arrangements between institutions allowing the IRB of one institution to rely on the IRB of another institution for review of human subjects research. Investigators working at multiple institutions, each having an IRB, may choose to have one IRB become the IRB of record over some or all participating sites. This means that the AU IRB is either the reviewing IRB (IRB of Record) or is relying on another IRB for IRB oversight of the research activity.*

*If the study will utilize a reliance agreement or a single IRB, please describe which institution(s) will be relying on another IRB for review and which institution will be responsible for the IRB oversight of the relying IRB(s).*

*Note: Requests for Reliance Agreements should be submitted by the Augusta University study team. All request for Reliance Agreements should be submitted through IRB Net following the procedures outlined in Forms and Templates*

Click here to enter text.

8. Procedures Involved

a. Describe the procedures involved to include those procedures that are standard evaluation and/or care and those that are solely for research purposes:

For research purposes:

*Following consent*– Block randomization to the twice daily irrigation with alkalinization or betadine will be done by random number generator. Out of the list of positives for the day (from both the lab and any call ins) we will call in sequential order to obtain a consecutive sample that meets all inclusion criteria and do not meet any exclusion criteria.

For the other patients on the list from the lab who tested positive for COVID-19 and meet screening criteria (age 55 or older, etc.) their information will be kept on a password protected spreadsheet and stored on the _HUMAN folder in box (also referred to as a research box) so that approximately one month later we can go back and determine hospitalization rates to be compared to the hospitalization rate of the study group.

*Following consent participants will have a box of study supplies* delivered to their residence by a member of the research team using COVID-19 precautions (masks, maintaining 6 ft. or more physical distance, door drop off). At this time the research team member will ask the participant to confirm receipt of the materials via text.

Either NeilMed nasal irrigation unit or Navage will be available on different days, with a goal of 50% of each device used in order to avoid concerns about conflict of interest.

Participants will be called (Day 2 call) after they receive their materials to answer any questions. Directions for nasal lavage will be included with the materials and via email. Participants will be instructed to watch a 1 minute video https://www.youtube.com/watch?v=DYZDeiOVJx0 on days with NeilMed supplies, or

<https://www.youtube.com/watch?v=qIfgNNLX1L8> if they received a Navage.

*Intervention – twice-a-day nasal lavage*

*Twice-a-day virucidal group:* Participants randomized to betadine will receive 2 gallon jugs of distilled water, two NeilMed Sinus Irrigation bottles and 28 salination packets (with some extras), OR one Navage unit with 28 SaltPods (and some extras), and a cardboard receptacle labeled “used saline containers” to keep track of adherence.

Those randomized to receive betadine will also receive one bottle of povidone-iodine, a one-sheet instruction with photographs demonstrating how to add ½ tsp betadine in addition to the salination packet to the sinus irrigation bottle or Navage unit reservoir prior to SaltPod, along with a ½ tsp measuring spoon.

*Twice-a-day alkalinized group:* Participants randomized to alkalinization will receive 2 gallon jugs of distilled water, two Neilmed bottles with 28 saline packets, OR one Navage unit with 28 SaltPods, and a cardboard receptacle labeled “used saline containers” to keep track of adherence. Those randomized to alkalinization will also receive a box of baking soda, ½ tsp measuring spoon and instructions on how to add the baking soda.

When study materials are procured the identifying information for the products (lot number, etc.) will be recorded so that we can track what materials were given to the participants. We will use the accountability log and the device accountability log (see attached).

As part of the materials box an instruction sheet will be included (see attached) that will detail the process for nasal irrigation and equipment cleaning. The websites for on-line resources will be included as well as study team contact information. Participants will be reminded on the instruction sheet that they should be the only one using the equipment for their safety and the safety of other household members. Information on this sheet will also include procedures for tracking material use via Qualtrics (see data collection sheet) and remind participants that at the end of the study they will keep or discard of all used and unused materials but that none are to be returned to researchers nor transferred to another person.

Prompts to complete study materials (see attached) will be sent to participants (text and/or email) via Qualtrics twice a day for the duration of the study period.

- Measures 6.1, 6.2, 6.3 (below) will be collected one time on enrollment.
- Measures 6.4, 6.5 will be collected in daily log on paper or online survey

Measure 6.6 will be collected at study conclusion after study day 28. If hospitalization occurred we will gather this information from The Georgia Regional Academic Community Health Information Exchange (GRAChIE) which serves healthcare providers across Georgia bringing health information from one healthcare professional to another, seamlessly connecting health information. GRAChIE health information exchange allows healthcare professionals to appropriately access and securely share a patient’s health information electronically through a member’s EMR system.

6.1 *Background information*. Patients will complete a demographic questionnaire that includes age, date of birth, residence type (homeless, single family, apartment living or group living), county of residence, sex (male, female, unknown, other), race, and ethnicity. (See data collection instrument.)
6.2 *Patient medical history.* Patient will be asked preexisting medical history as found on the CDC person of interest form, including Chronic Lung disease (Emphysema, COPD), Asthma, Type 1 Diabetes Mellitus, Type 2 Diabetes Mellitus, Cardiovascular Disease, Hypertension, Chronic Renal Disease, Weight and height to calculate obesity with BMI>30, Immunocompromised condition (if YES specify medications they are taking), Current cancer, Current Smoker, treatment if any, and symptoms. (See data collection instrument.)
6.3 *Interventions:* Patients will be asked if they have used any interventions against COVID, including acetaminophen, ibuprofen, DOAN’s Packets, aspirin, Melatonin, Vitamin C, Vitamin D, Quercetin, Zinc, Chinese Traditional Medicines, or other. (See data collection instrument.)
6.4 *Current Symptoms.* Patient will be asked for number of days since first subjectively sick, loss of smell, loss of taste, fatigue, presence or absence of fever >100.4 (if yes how many days), subjective fever, chills, muscle aches, runny nose, cough (new onset or worsening of chronic cough), shortness of breath, nausea or vomiting, headache, abdominal pain, diarrhea (>3 loose/looser than normal stools/24 hours). (See data collection instrument.)

*6.5 Daily Adherence:* Participants will receive two daily reminders to do nasal irrigation and subsequent verification, and asked to keep a daily study diary using Qualtrics. The additive packages for nasal irrigation will be attached to a one page card with numbers and days to help patients visually assess whether they have done it yet that day. During the daily verification participants will be asked to report from their diary the presence or absence of the following symptoms: loss of smell, fatigue, cough, muscle aches, harder to breathe, AND to report any other medications taken, AND to provide a photograph of this card as well as the number of used packets in the box. (See data collection instrument.)

An investigator will call the patient or their designated contact at Day 2, 7, 14, and 28.

*6.6 Outcomes*: The following outcomes will be collected via GRAChIE as described above (see data collection spreadsheet): visit to an emergency department Y/N; chief complaint was the patient hospitalized y/n (if yes admission date/discharge date); was the patient administered supplemental oxygen y/n was the patient admitted to an intensive care unit y/n was patient mechanically intubated Y/N was patient given assisted ventilation with CPAP/HighFlow/BIPAP Y/N did the patient die as a result of this illness Y/N (date of death).

For the purposes of the study, COVID-19 related chief complaints will include any of the top five symptoms associated with emergency department presentation: Cough, Fever (subjective or >100.4°F/38°C), Myalgia, Headache, or Dyspnea.^18^ Other ED presentations resulting in discharge without subsequent admission will not meet the criteria of morbidity severity related to this study. In cases where patients are admitted, discharge diagnosis or cause or contributing cause of death will be considered for categorization of COVID-related or not-COVID related. In cases where potential for COVID19 related admission or death is ambiguous, a member of the research team (MD) will be asked to assess the information gathered from the chart and make a determination of COVID or non-COVID related.

18. MMWR Morb Mortal Wkly Rep. 2020;69(24):759

8.1. Participants will receive two prompts a day to complete the nasal lavage and complete the data collection diary.
8.2. Participants who opt for the online data collection tool will be provided with a phone call to follow up on data collection when they don’t complete online data collection for two days. In the case of a patient reporting a summary number of irrigation events or a range of irrigation events, the estimate or median number of irrigation events range will be used for analysis. If no data is obtained after confirmation of the initial irrigation, the patient’s data will be excluded from the continuous variable irrigation analyses, but will be included in analysis of betadine/alkaliniaztion and Navage/Neilmed by intention to treat.
8.3. Sources of Research Material – See “measures”. Background information will be obtained from the patient at the time of enrollment. (see Measures). Outcomes will be assessed by records stored in GRAChIE and from the participant and designated contact person.
8.4. Instrument - For the study, an irrigation diary will be created, and packets of nasal irrigation will be attached to a one page card with numbers and days to help patients visually assess whether they have done it yet that day. A photograph of this card will be requested as part of the patient diary.
8.5. Follow-up – An investigator blinded to allocation will call the patient on Day2, Day 7, Day 14 and Day 28 to check on their current symptoms (if the participant is unable to be contacted after three attempts on any of these designated days the secondary contact will be called up to three times to check on the participant's current symptoms.

For the control group, after enrollment is stopped, eligible patients with a positive test who were not enrolled will be separated into two groups: those who refused enrollment and those who were not able to be reached or responded too late to be enrolled on the day of contact. One control will be chosen from each group by the following criteria: sex, race, age +/- 2 years, date of test +/- 1 week. If there are no matches following this schema, date of test will be expanded to +/- two weeks. If race is listed as “unspecified”, initial experience with enrolled patients supports that commonly this is chosen by Black patients. Thus, when matching with a Black patient, two “unspecified” matches will be chosen and searched in order in GRAChIE. The first patient with matching race will be the Control included. If the criteria for matching cannot be met with 1 from each of the “refused” and “uncontacted” pools, two matches will be found in the other pool. The EMR will then be searched for the following information from the Control group within 28 days of the day they could have been enrolled (see data collection spreadsheet) : visit to an emergency department Y/N, chief complaint; was the patient hospitalized y/n (if yes admission date/discharge date); was the patient administered supplemental oxygen y/n); was the patient admitted to an intensive care unit y/n was patient mechanically intubated Y/N); was patient given assisted ventilation with CPAP/HighFlow/BIPAP Y/N did the patient die as a result of this illness Y/N (date of death).

Click here to enter text.

b. Describe and explain the study design:
   If the study involves multiple conditions where each condition involves different procedures, please provide a table that breaks down the procedures by condition and in chronological order. Include when and where they are performed.

This is a single center randomized controlled non-blinded clinical efficacy study evaluating nasal irrigation with either alkalinized saline or dilute Povidone-Iodine twice a day for 14 days for the reduction of COVID-19 severity in COVID+ outpatients.

Click here to enter text.

c. Data Types and Source Records:

*Briefly describe the actual source records or measures that will be used to collect data about participants. (All surveys, interview scripts, and data collection forms will be attached elsewhere in the application. Do not add other documents to the protocol.) Describe what data will be collected and how it will be collected at all measurement/data collection time-points.*

d. Describe the procedures performed to lessen the probability or magnitude of risks:

Data will be collected via Qualtrics, phone calls, and GRAChIE. Information will be stored and collected in a coded manner. Data will be stored in the _HUMAN folder in Box (also referred to as a research box) for secure storage. A master code list will be used to assign a study number to each participant.

In addition, we will be following all CDC guidelines and Augusta University research policies in the collection of this data. We have designed the study to use existing infrastructure possible to recruit for the study and will use masks and social distancing for no-contact study material delivery by a member of the research team.

Clk hereto enter text.

e. Describe the duration of an individual subject’s participation in the study and the time involved also include the overall duration of the project:

A participant’s involvement in the study will last 28 days from the time of enrollment. For the first 14 days approximately 15-30 minutes will be involved in order to complete both nasal lavages as well as to provide information in the daily diary, and respond to prompts or phone calls.

Click here to enter text.

9. Data and Specimen Management

| a. *Describe the data analysis plan, including any statistical procedures:*   Interim analysis planned at 200 patients. In the event that the patients enrolled show an overwhelming trend toward less or greater severity or lack of efficacy with povidone-iodine versus alkalination for safety or for preponderance of efficacy of irrigation itself, study will be halted to analyze the case: control outcomes for disease severity: ED visit, admission, ICU, death, and historic outcomes for household transmission. The povidone-iodine/alkalinization and frequency of irrigation endpoints will be evaluated separately as power permits.   8.6. *Endpoints*      8.6.1. Primary – Primary outcome will be a statistical difference between need for hospitalization visit by number of nasal irrigations completed and by group. The case:control analysis will be conducted by intention to treat.      8.6.2. Secondary - Secondary outcomes include differences between symptoms, supplemental oxygen use, ICU admission, mortality.     Statistical Analysis - Initial analyses will be conducted to determine whether demographic variables are related to the following dependent variables: hospital visit; oxygen use; admission; ICU admission; mortality. To examine sex and racial differences on the dependent variables, *t*-tests will be used unless non-normal distribution is noted, in which case Mann Whitney U will be used. Baseline measures of duration, pre-existing conditions, and symptoms will be compared across the conditions to ensure that the randomization process was effective. Analyses of compliance by device and the previously mentioned dependent variables will be conducted to determine whether the brand of device impacted outcomes. If any of these analyses result in significant findings, they will be considered as covariates in subsequent analyses. To test the effect of treatment change in disease severity, either t-tests for normally distributed data or the Mann-Whitney will be used. Pearson’s correlation coefficient will assess if an inverse linear relationship exists between number of irrigation uses and illness severity outcomes.   9.2.1 Based on histogram plotting of the main outcome variable, hospital use, it will be determined whether *t-*tests for normally distributed data or the Mann Whitney will be used to test for differences between BID and once daily conditions. Separate analyses will be conducted examining preexisting conditions, self-administered interventions, and duration of symptoms. Paired t-tests will evaluate the effect of BID alkalinized rinses on hospital use, oxygen, ICU, and death. Given previously established correlation between the preexisting conditions and disease severity, Tukey’s procedure will be used to control for Type I error rather than the more conservative Bonferroni correction.   Click here to enter text. | **☐** N/A |
| --- | --- |
| b. When applicable, provide a power analysis:    9.3 *Analysis*   *Power Analysis:* The analysis will include treatment compliance with doses as a continuous variable correlated with clinical outcome), and full compliance (two rinses daily until resolution of symptoms or hospitalization). A 2-sided Fisher’s exact test for binary outcomes will be used, with an alpha of 0.05 and beta of 0.80 requiring 100 patients in each group, anticipating an a priori severity of 20% needing oxygen or hospitalization and a 50% reduction to 10% being significant. Subanalyses will evaluate the prevalence of relevant pre-existing conditions and severity of symptoms at initiation of treatment.   9.3.1 Below is a table of different effect sizes and the associated sample size assuming a statistical power of 80%.   Effect size n per group Total N  .3  .5  .7  Different effect sizes have been noted for different nasal irrigation studies, with most starting within 48 hours of influenza or rhinoviridae with shorter incubation periods.  9.3.2 Case Control matching: Power was calculated two ways, by running a simulation comparing the likelihood of hospitalization, and using a z-test comparing independent proportions. Both analyses assumed reduced variation in proportions due to accounting for the correlates of age, sex, and race, and resulted in similar calculations. Assuming that 25% of the matched control participants and 10% of the treatment participants would visit the hospital (60% reduction), 79 participants in the nasal irrigation condition would have .80 (80%) power to detect that proportional difference. The inability to evaluate and match underlying conditions in the control group can increase variability, thus a 1:2 matching schema was chosen recommended over 1:1.  Click here to enter text. | **☐** N/A |
| c. *Describe how data and specimens will be handled:* | **☐** N/A |
| i. What information will be included in that data or associated with the specimens?   A Master code list will be used to assign a study identification number to each participant. The participant number will be associated with any data that are collected so the data will be stored with no directly linked identifiers. An additional code list will be used to connect the subject ID with the ID assigned for the chart review portion of the study. | |
| ii. Where and how data and/or specimens will be stored?   _HUMAN folder in box (also referred to as a research box) | |
| iii. How long will the data and/or specimens be stored?   3 years after end of study | |
| iv. Who will have access to the data or specimens?   Research Team | |
| v. Who is responsible for receipt or transmission of the data and/or specimens?   Research Team | |
| vi. How will data and/or specimens be transported?   Electronically | |

10. Provisions to Monitor the Data to Ensure the Safety of Subjects ☐ X N/A

*The plan might include establishing a data monitoring committee and a plan for reporting data monitoring committee findings to the IRB and the sponsor.*

a. Describe the plan to periodically evaluate the data collected regarding both harms and benefits to determine whether subjects remain safe.

Click here to enter text.

b. Describe what data are reviewed, including safety data, untoward events, and efficacy data.

Click here to enter text.

c. Describe how the safety information will be collected (e.g., with case report forms, at study visits, by telephone calls with participants).

Click here to enter text.

d. Describe the frequency of data collection, including when safety data collection starts.

Click here to enter text.

e. Describe who will review the data.

Click here to enter text.

f. Describe the frequency or periodicity of review of cumulative data.

Click here to enter text.

g. Describe any conditions that trigger an immediate suspension of the research.

Click here to enter text.

11. Withdrawal of Subjects	☐ N/A

| a. *If applicable, describe anticipated circumstances under which subjects will be withdrawn from the research without their consent.*   Withdraw from study: Participants can end participation in the study at any time. A subject’s participation in the study will be discontinued for any of the following complications, but will still be included in the final analysis:   - inability to tolerate irrigation - inability to perform irrigation - The sponsor or study doctor decides to stop the study. - The study doctor stops your taking part in the study for your safety. - You are not eligible to take part in the study. - Your condition changes and you need treatment that is not allowed while you are taking part in the study. - You do not follow the instructions from the study staff.   Click here to enter text. | **☐** N/A |
| --- | --- |
| b. If applicable, describe any procedures for orderly termination.   Stopping Rules. The study will be temporarily suspended, if a single patient has evidence of any adverse effects of irrigation.  Click here to enter text. | **☐** N/A |
| c. If applicable, describe procedures that will be followed when subjects withdraw from the research, including partial withdrawal from procedures with continued data collection.   If a participant withdraws or is withdrawn any data collected up to that point will be kept and used in the analysis as possible.  Click here to enter text. | **☐** N/A |

12. Risks to Subjects

| a. *List the reasonably foreseeable risks.*    8.7. *Potential Risks -* The possibility exists that inability to complete nasal irrigation could cause coughing or that the splashing of the nasal irrigant itself could increasing exposure in households. There is a possible risk of infection due to improper cleaning of the irrigation devices. It is known that mild itching/stinging with the rinse is a possible side effect of the treatments. This will be included in the ICD and explained to participants during the consent process. There is the possibility of a loss of confidentiality but procedures have been put in place to prevent this.   Click here to enter text. | |
| --- | --- |
| b. If applicable, describe any costs that subjects may be responsible for because of participation in the research.   Click here to enter text. | **X ☐** N/A |
| c. If applicable, describe risks to others who are not subjects.   Click here to enter text. | **X ☐** N/A |

13. Potential Benefits to Subjects

*Describe the potential benefits that individual subjects may experience from taking part in the research.*

There is the potential for a direct and even life-saving benefit to the subjects who volunteer to participate.

14. Confidentiality

*Describe the procedures for maintenance of confidentiality.*

Any information that is obtained in connection with research that can be identified with a subject will remain confidential. Patients will be assigned a study number which will match their informed consent, and all information will be recorded and stored in a coded manner. Statisticians involved in the project will have access to de-identified data for the purposes of analysis. Copies of the informed consents will be kept by the AU Department of Emergency Medicine. Patient samples will not be obtained. Augusta University and The Institutional Review Board reserve the right to audit any research files to assure the quality of any data used in this research.

Click here to enter text.

15. Incomplete Disclosure, Authorized Deception, or Deception	☐ X N/A

*See the Deception Policy on Augusta University Website.*

*If the study will use incomplete disclosure or deception, describe the incomplete disclosure or deception, and provide a rationale explaining why it is necessary to the research.*

*Describe the debriefing process that will be used to make participants aware of the incomplete disclosure or deception, including their right to withdraw any record of their participation.*

Click here to enter text.

16. Consent Process

*If you are obtaining consent of subjects describe the consenting process. Be sure to include the process to be used if enrolling illiterate, non-English speakers, individuals with impaired decision making capacity to consent, as applicable.*

Eligible patients who are obtained from the list of positives provided by the testing lab will be contacted by phone. The study will be explained to them and any questions they have will be answered. If they choose to participate remote consent will be obtained and documented by a member of the research team.

Remote consent will be obtained using the Remote Informed Consent Process that is included in the Phone Script for Active Participant Group 1 and the Phone Script for Active Participant Group 2 (see attached documents). For those with access to email or fax the consent document will be provided that way and the rest of the steps followed.

Because of the time sensitive nature of the study any participant who does not have access to email for fax will be provided a copy of the entire consent document as the first step in the door drop off. After dropping off the document the researcher will return to their car and call the participant as well as adding a 3^rd^ party witness to the process and follow the remote consent procedures process as described for those participants who do not have access to a fax or email.

17. Compensation for Research-Related Injury

*This section is not required when research involves no more than Minimal Risk to subjects.*☐ X N/A

a. Describe the available compensation in the event of research related injury.

Click here to enter text.

18. Qualifications to Conduct Research and Resources Available

*Describe the qualifications of you and your staff to conduct this research. The IRB is looking for information such as area(s) of expertise, past research experience, relevant certifications, etc.*

*For international research or research with vulnerable populations, describe the qualifications (e.g., training, experience, oversight) of you and your staff as required to conduct the research. When applicable describe the knowledge of the local study sites, culture, and society. Provide enough information so the IRB knows that you have qualified staff for the proposed research.*

*Note: If you specify a person by name, a change to that person will require prior approval by the IRB. If you specify people by role (e.g., coordinator, research assistant, Sub-Investigator, or pharmacist), a change to that person will not require prior approval by the IRB, provided that person meets the qualifications described above to fulfill their roles.*

Matt Lyon, MD is a professor of Emergency Medicine, Vice Chairman for Academic Programs & Research for the Department of Emergency Medicine and also the Director of the Center for Ultrasound Education for Augusta University. Recently Dr. Lyon has been helping to lead AU's efforts in combating COVID-19 via telemedicine. Dr. Lyon has more than 20 years of research experience and has been the PI on more than 30 approved studies. Dr. Lyon will be supported by the Department of Emergency Medicine research team which includes Dr. Gibson who has years of research experience and currently serves as a Vice Chair for the IRB, Dr. Kuchinski who has been managing research for more than 3 years as well as Houlton Boomer who will be assisting with the regulatory. Any other study members will be trained in the protocol by the PI and research staff.

a. Describe the availability of medical or psychological resources that subjects might need as a result of an anticipated consequences of the human research.

N/A

b. Describe your process to ensure that all persons assisting with the research are adequately informed about the protocol, the research procedures, and their duties and functions.

The PI and members of the research staff will ensure that all member of the study team are trained in the protocol.

.
